# Tracing and testing the COVID-19 contact chain: cost-benefit tradeoffs

**DOI:** 10.1101/2020.10.01.20205047

**Authors:** Jungyeol Kim, Xingran Chen, Shirin Saeedi Bidokhti, Saswati Sarkar

## Abstract

Traditional contact tracing for COVID-19 tests the direct contacts of those who test positive even if the contacts do not show any symptom. But, why should the testing stop at direct contacts, and not test secondary, tertiary contacts or even contacts further down? The question arises because by the time an infected individual is tested the infection starting from him may have infected a chain of individuals. One deterrent in testing long chains of individuals right away may be that it substantially increases the testing load, or does it? We investigate the costs and benefits of testing the contact chain of an individual who tests positive. For this investigation, we utilize multiple human contact networks, spanning two real-world data sets of spatio-temporal records of human presence over certain periods of time, as also networks of a classical synthetic variety. Over the diverse set of contact patterns, we discover that testing the contact chain can both substantially reduce over time both the cumulative infection count and the testing load. We consider elements of human behavior that enhance the spread of the disease and lower the efficacy of testing strategies, and show that testing the contact chain enhances the resilience to adverse impacts of these elements. We also discover a phenomenon of diminishing return beyond a threshold value on the depth of the chain to be tested in one go, the threshold then provides the most desirable tradeoff between benefit in terms of reducing the cumulative infection count, enhancing resilience to adverse impacts of human behavior, and cost in terms of increasing the testing load.

---

To slow down the spread of COVID-19, public health authorities like the US Center for Disease Control and Prevention (CDC) have recommended to test those who have in the recent past been in physical proximity with a patient who has tested positive, even when the contacts do not exhibit any symptom [30]. This preemptive action, commonly known as contact tracing, is deployed because given how contagious the disease is, a patient is likely to have passed on the contagion to his contacts, and the infected contacts have the potential to infect others even before they show symptoms [12]. Moreover, the CDC estimates that up to 70% percent of the infected individuals are asymptomatic, showing no symptom throughout the entire course of the disease [25], and clinical research has revealed that the asymptomatic individuals can infect others [16, 29]. Testing and isolating the infected can stop these infected individuals from spreading the disease early on, that is, while they do not show symptoms, as compared to the strategy that tests only those who show symptoms and seek medical help. Slowing down the spread by testing the contacts comes at the cost of an increase in the testing load as compared to the latter policy, yet, the cost-benefit tradeoff for contact tracing is understood to be substantially favorable (cost is the testing load, benefit is the ability to contain the outbreak).

A question that naturally arises is if cost-benefit tradeoffs may be enhanced through generalizations of the core concept of contact tracing - this is what we seek to answer in this paper. In the time that elapses between when an individual is infected and until he is tested, the disease spreads from him through a chain of several hops - he infects those he is in contact with, those he infects infect their contacts, the infected contacts infect their contacts, and so on. Fewer people are likely to be infected if we preemptively test not only the direct contacts of an individual who tests positive, but contacts of the contacts and so on. Such testing will enable us to identify and isolate the individuals further down the chain who have imbibed the disease, earlier than if we had tested only the direct contacts of those who have tested positive and reached down the chain progressively. Earlier isolation of the infected reduces the number they infect. Does such aggressive preemptive testing schemes necessarily increase the overall number of tests? The answer is not apriori clear as reduction in overall infection spread through such a testing strategy may eventually reduce the number of tests required, as illustrated in Figure 1.

**Figure 1:**
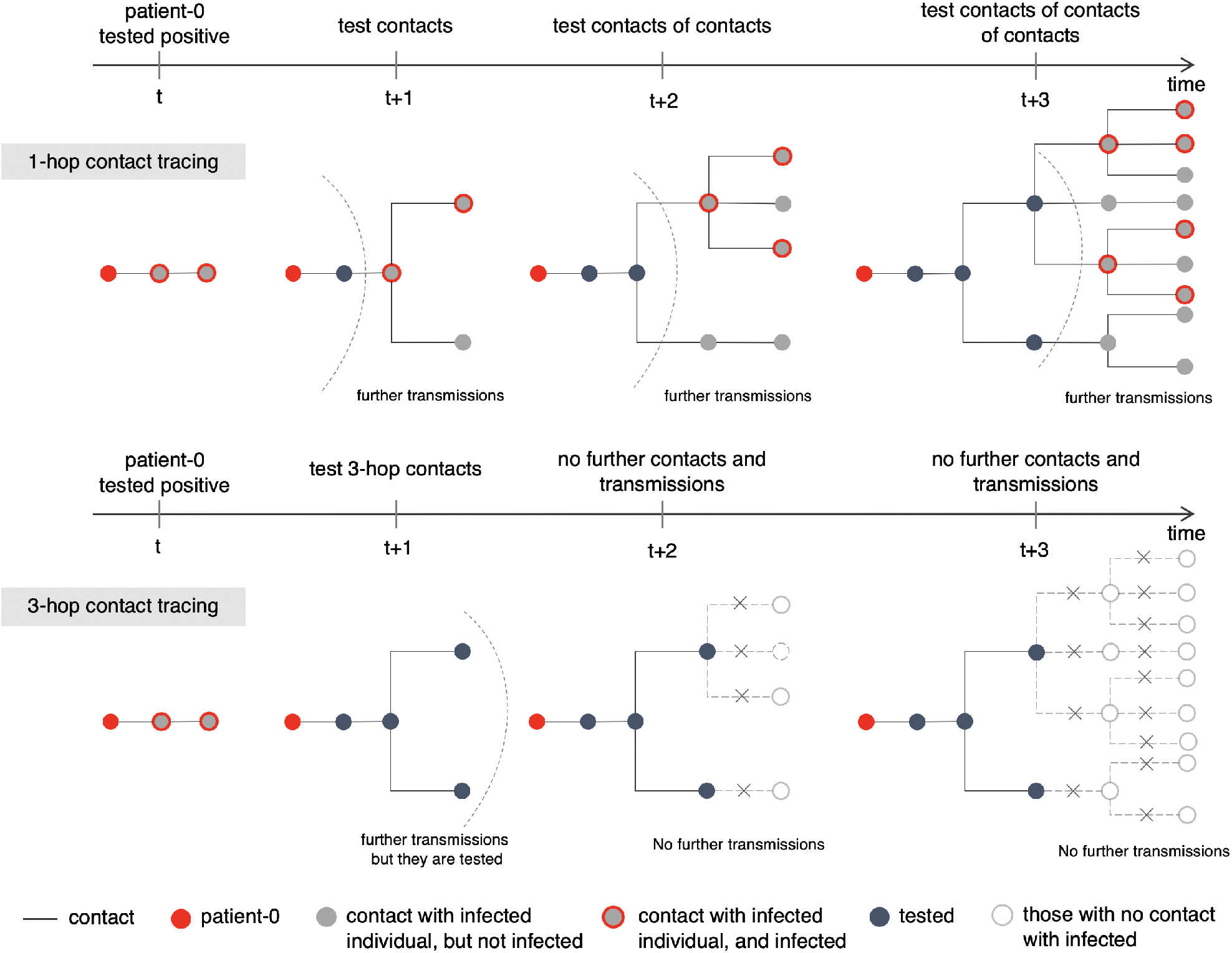
Illustration of multi-hop contact tracing. Illustration of 1-hop contact tracing (i.e., testing only the direct contacts of those who test positive) and 3-hop contact tracing (i.e., testing the direct, secondary and tertiary contacts of those who test positive). The time at which a health authority tests the patient-0 (red) was after the infection has propagated 2 hops. By *t* +3 time units, both testing policies test 4 individuals (black) other than the patient-0; the 3-hop policy tests and isolates the positive ones in a shorter time, while 1-hop tests and isolates them progressively and therefore over longer times. Accordingly, only 3 individuals are infected under the 3-hop policy, while 10 individuals are infected under the 1-hop policy.

We formalize this aggressive preemptive testing scheme as *k-hop contact tracing* (*k*), where *k* = 0 does not trace contacts and tests only those who show symptoms and seek medical help, *k* = 1 is the traditional contact tracing that tests the direct contacts of an individual who tests positive, *k* = 2 additionally tests the contacts of the contacts, *k* = 3 tests yet another hop of contacts, and so on. Our investigation will quantify the 1) benefits i.e., reduction in the number of individuals infected over time, 2) costs, i.e., increase in total number of tests, with increase in *k*, for a wide variety of disease parameters, contact patterns, extent of willingness of individuals to cooperate with the health officials on testing. We investigate for a wide variety of the above parameters because values of the parameters that arise in practice are not definitively known at this nascent stage of research on the novel disease, and will in general be different for different ambiences. The goal of our investigation is to reveal if this natural generalization has any merit and provide specific policy recommendations with respect to testing strategies. We will also examine whether there arises the principle of diminishing return, that is, increasing the number of hops beyond a certain *threshold* only marginally decreases the infection count but noticeably increases the testing load - if so, such threshold, which we seek to identify, provides the optimum cost-benefit tradeoff.

Human behavior plays an important role in determining the nature and extent of the spread of an infectious disease and the efficacy of tests:

- When a contagious individual interacts with a susceptible individual, the latter is infected with a probability whose value depends on various factors such as the duration, environment (e.g., indoor or outdoor) of the interaction, distance between interacting individuals, protective gears worn by the individuals involved, personal hygiene like hand washing observed right after interactions etc. [28]. The probability is understood to be higher for longer durations, shorter distances, indoor environments and lower when appropriate protective gears are worn and good personal hygiene is observed [7, 26, 27]. But, ambiences that reduce probability of infection can not always be guaranteed owing to professional necessities and voluntary choices. For example, several societal interactions will have to be in indoor environments (e.g., among customers and staff of indoor businesses, fellow travelers in elevators, public transports), for long durations (inside business units, public transport, elevators), in close proximity (inside elevators, public transport). Protective gears like masks and personal hygiene like hand washing can be recommended but not enforced. The most effective masks like N95 masks may not be available for every individual and may also be inconvenient for daily use for common people, masks may not always be worn properly. The net effect of all of these is that probability of infection can not be controlled and may be high in various circumstances.
- Human behavior also affects the reach of testing. It has been documented that only a fraction of contacts of patients who test positive can be tested, as many contacts either do not consent or can not be traced [14]; we refer to this fraction as *cooperativity*. Many contacts for example do not respond to outreach by public health authorities. Contact-tracing apps installed in wearable or hand-held devices can temporarily record contacts between individuals and communicate pertinent information to the centralized database once an individual tests positive [9]. Such contact tracing apps have been rolled out in Germany, Singapore, England and some states in the US. These increase the fraction of cooperativity, but still do not guarantee full cooperativity as individuals may function in areas of poor signal and may not carry their personal devices all the time. Besides downloading such apps can not be mandated owing to concerns on privacy and considerations of civil liberties and free choice. Thus their acceptance to the populace is unclear at this point. Hence cooperativity may be low in real life.

Resilience of a testing strategy to elements of human behavior that enhance the spread of the disease and lower the efficacy (i.e., reach) of testing constitutes an important benefit. We refer to *k*-hop contact testing for *k* > 1 as *multi-hop contact tracing* and ask if and in what sense multi-hop contact tracing enhances resilience compared to 1-hop contact tracing (that is, the traditional contact tracing), that is, can multi-hop contact tracing somehow offset the larger spread due to imponderables of human behavior?

The significance of our investigation also draws from the fact that the testing recommendations by the regulating authorities have not been finalized yet, and recommendations are being continually adapted as new facets are being discovered. For example, in the last few months the CDC has changed its recommendation about testing the asymptomatic individuals multiple times [29]. Thus, new findings about testing strategies are likely to find their way to practice now with fewer legacy-related complications.

We consider a discrete time stochastic evolution of COVID-19 in a population that initially consists of susceptible and a few contagious individuals. We model the progression of the disease using a compartmental model (Figure 2). The disease spreads from the contagious to the susceptible individuals through mutual interaction. In any given interaction with a contagious individual, a susceptible is infected with a probability. After a *latency period* (the presymptomatic-latent and asymptomatic-latent are in this latency period), the newly infected individuals become contagious. Specifically, at the end of the latency period, the individuals either become *presymptomatic* (the stage before exhibiting symptoms), or *asymptomatic* (that is, they never show symptoms). Presymptomatics proceed to become *symptomatics* in the next stage. Presymptomatics, asymptomatics, symptomatics all however are contagious.

**Figure 2:**
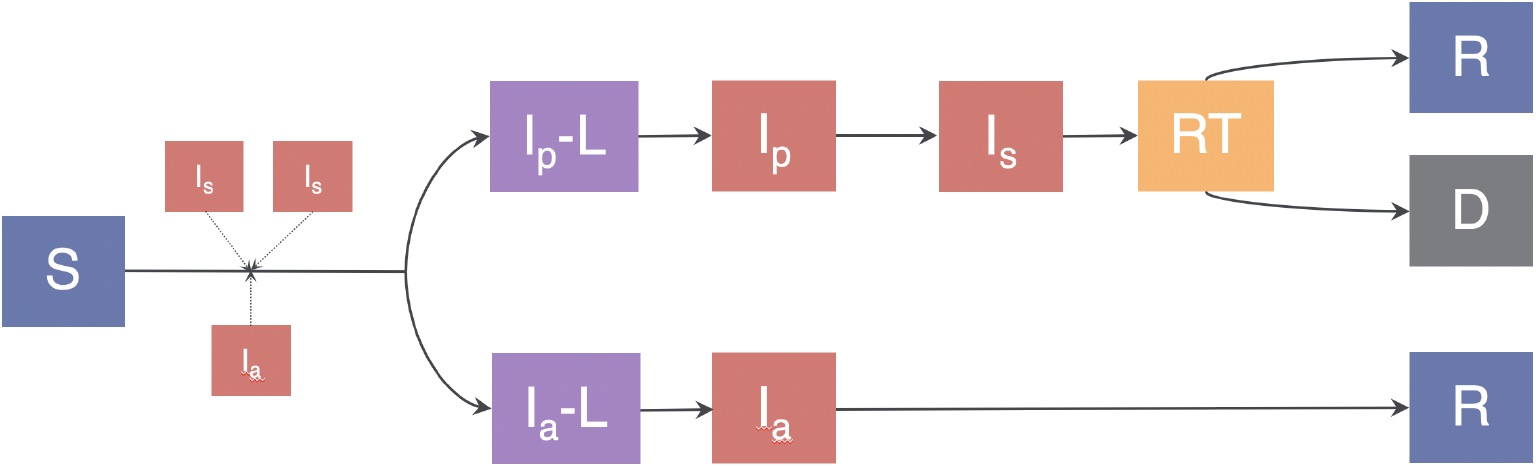
Virus transmission model illustration. The Compartmental model consists of the following compartments: Susceptible (*S*), Presymptomatic-Latent (*I*_*p*_-*L*), Presymptomatic (*I*_*p*_), Symptomatic (*I*_*s*_), Ready-to-Test (*RT*), Asymptomatic-Latent (*I*_*a*_-*L*), Asymptomatic (*I*_*a*_), Recovered (*R*), and Dead (*D*).

We assume that test results are obtained in the same day, owing to the availability of reliable RT-PCR and antigen tests that are able to do so [11, 23] (recent antigen test authorized by the FDA under an emergency use can give results in 15 minutes with 97.1% sensitivity).

Given the imponderables of human behavior, we consider a range of values for the parameters they affect directly, namely, cooperativity and probability of infection. For the former we consider the following values: 0.2, 0.5, 1.0. In addition, we assume that the contacts who can be identified are identified within a day, contact-tracing apps can provide such turnaround times and can be easily modified to trace and alert indirect exposures through iterative algorithms. Tests can then be scheduled the next day through the same apps. As to the probability of infection, we use this term to denote the probability with which a symptomatic individual infects a susceptible individual during a contact, and consider the following values: 0.04, 0.2, 0.4. We consider that a presymptomatic individual infects a susceptible with the same probability as a symptomatic individual, and that an asymptomatic individual infects a susceptible with 0.75 times this probability [25].

We have simulated *k*-hop contact tracing for *k* = 0, 1, 2, 3 to large values of *k* for contact patterns obtained from real-world data and various synthetic topologies. We consider two contact networks obtained from real-world data of: 1) physical proximity of university students obtained from their smartphones equipped with Bluetooth over 28 days, as part of the Copenhagen Networks Study [22] (*University student contact network*), and 2) locations and time-stamps of individuals in Tokyo, Japan which they provide over a social network (Foursquare) over 100 days (*Foursquare contact network*). These capture certain semblances of reality in that they are obtained from spatio-temporal records of actual human presence; here the contact patterns vary over time which is also what happens in reality. Yet, these are but constructed from only two sets of data, which may not be representative of all contact patterns. We therefore examine whether the phenomena observed in these also recur in some other very different networks, namely in two examples of classical synthetic networks: Erdős Rényi and scale-free networks. These two examples complement each other in some fundamental characteristics such as in the nature of the degree distribution. The degrees of the nodes represents the number of contacts of the corresponding individuals. The Erdős Rényi network is more regular, in that there is relatively low variance in the degree distribution. The scale-free network, on the other hand, has some nodes with high degrees (perhaps representing celebrities who interact with a large number of individuals) and many more nodes with low degrees (representing common folks); the degree distribution therefore has a high variance. For these the connections do not change over time, they are static in this sense. Any phenomenon that is observed in the four very different types of networks modeling human contact patterns is likely to recur extensively.

Refer to Methods for details on the systems we consider, the parameters we choose and further justification for the choices and the limitations thereof.

## Results

We start with a summary of our important findings from the simulations for *k*-hop contact tracing that we perform for *k* = 0, 1, 2, 3, …. We observe the following for multi-hop contact tracing (which is *k*-hop contact testing for *k* > 1):

- *Benefit of multi-hop contact tracing:* Multi-hop contact tracing considerably reduces the total number of infections over time compared to 1-hop contact tracing (that is, the traditional contact tracing). The reduction is higher with 1) increase in probability of infection 2) the decrease in cooperativity 3) decrease in the latency period. In addition, multi-hop contact tracing substantially reduces the average daily new infection count in the *peak infection period* (time until the daily new infection count peaks) compared to 1-hop contact tracing.
- *Cost of multi-hop contact tracing:* Over time the overall number of tests required in multi-hop contact tracing is usually *lower*, than that in 1-hop contact tracing. But, studying how the number of tests changes with time, initially the number of tests needed for multi-hop is somewhat (considerably in few instances) higher than for 1-hop contact tracing.

Increasing the value of *k* from *k* = 1, the number of infections and the number of tests over time decreases up to a threshold value of *k* (we observe that this threshold value is *k* = 2 or *k* = 3 in our simulations). Formally, we define the threshold point as the value of *k* at which the overall number of tests required (cost) is minimized. As *k* increases beyond this threshold value, we see a phenomenon of *diminishing return*, that is, the number of infections only marginally decreases and number of tests increases (considerably, in some instances). We substantiate the above findings with the results reported in Figures 3, 4, and 6; Figures 3 and 4 are for contact networks obtained from real-world data and Figure 6 is for synthetic topologies. In each case we report the average of 1000 simulation runs on given topologies.

**Figure 3:**
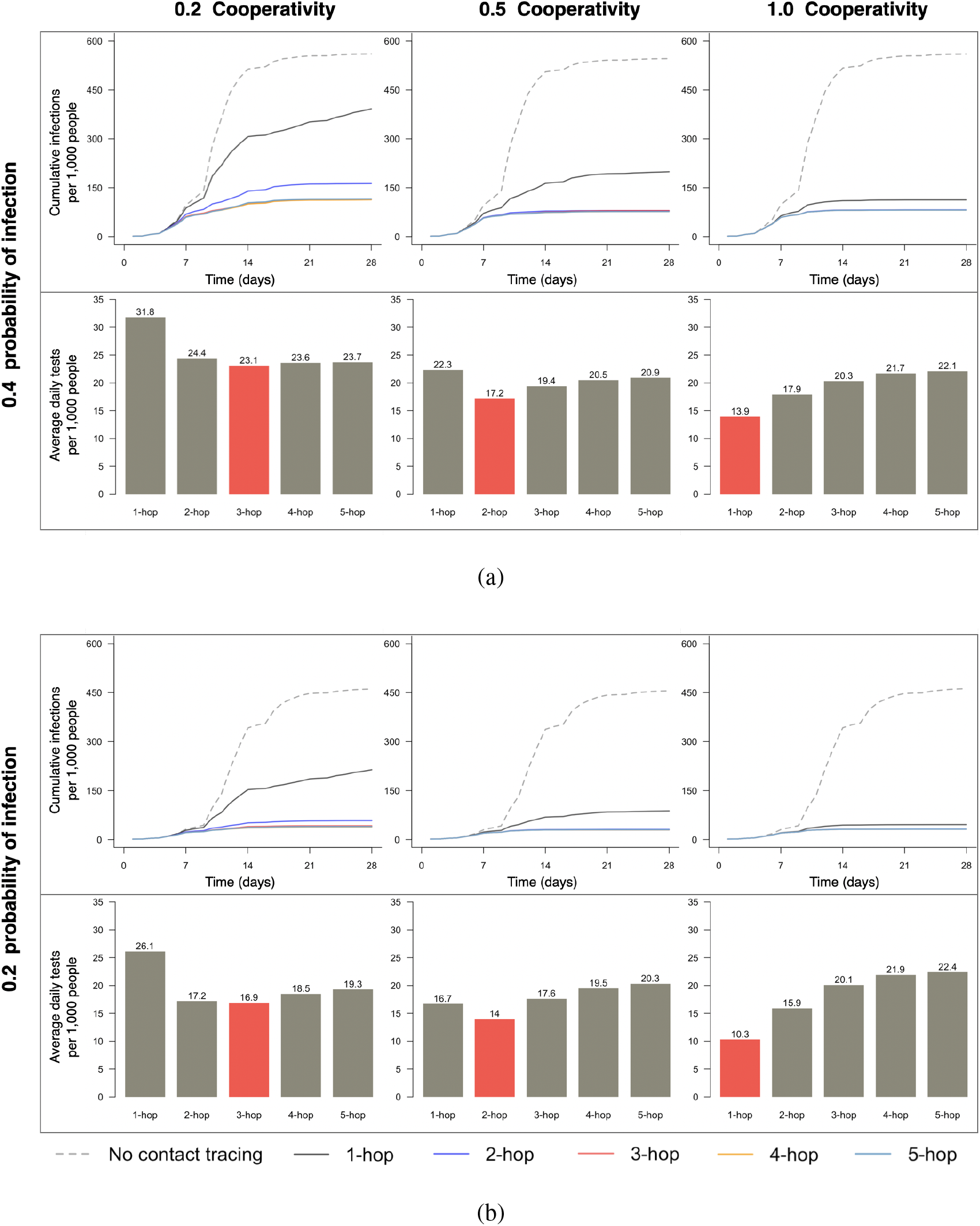
The cumulative number of infections and average daily tests required for *k*-hop contact tracing for various values of *k* for *University student contact network*. The red colored bar corresponds to the threshold value for *k*. For *k* exceeding the threshold value, the curves for the cumulative number of infections heavily overlap and become indistinguishable.

**Figure 4:**
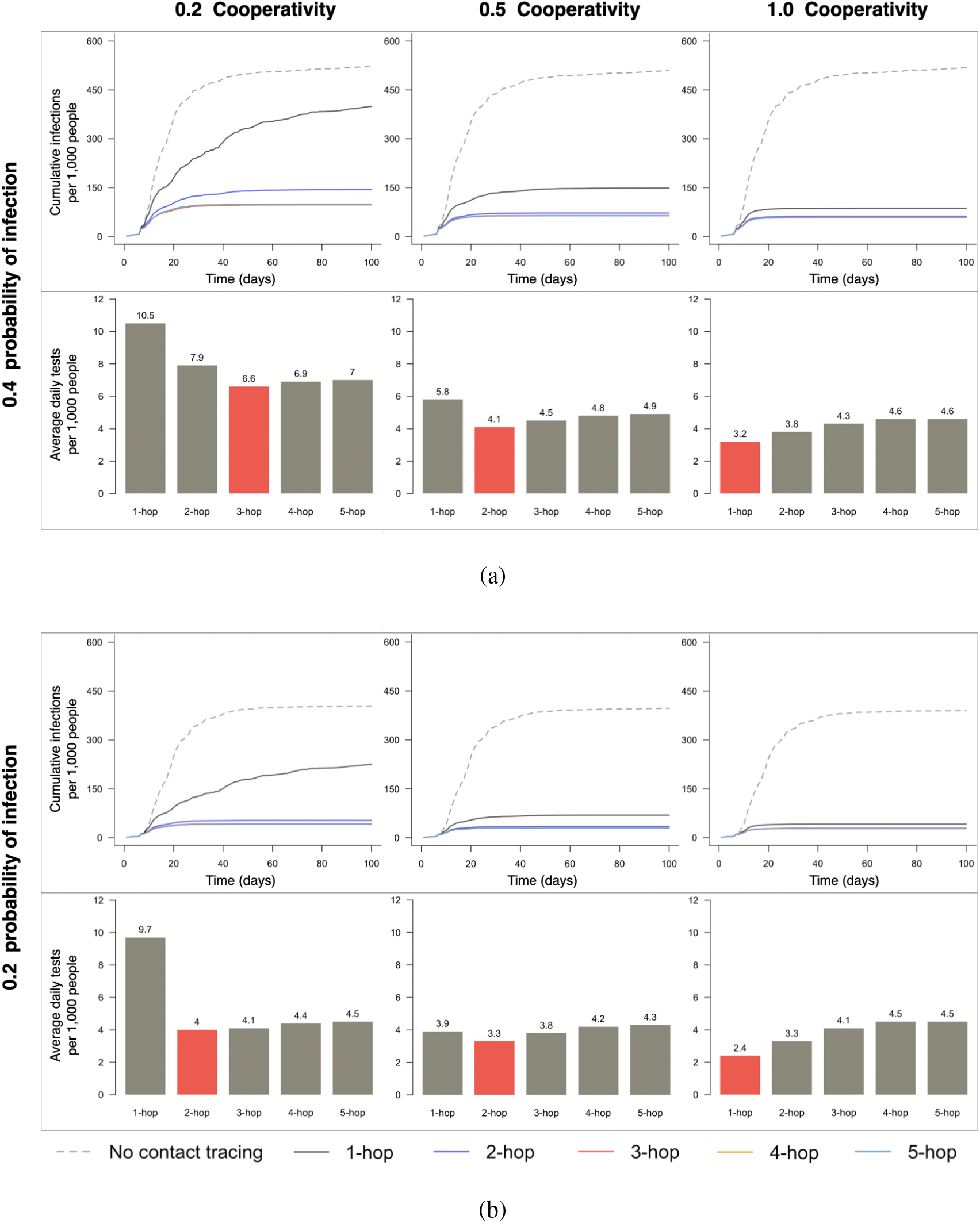
The cumulative number of infections and average daily tests required for *k*-hop contact tracing for various values of *k* for *Foursquare contact network*. The red colored bar corresponds to the threshold value for *k*. For *k* exceeding the threshold value, the curves for the cumulative number of infections heavily overlap and become indistinguishable.

**Figure 5:**
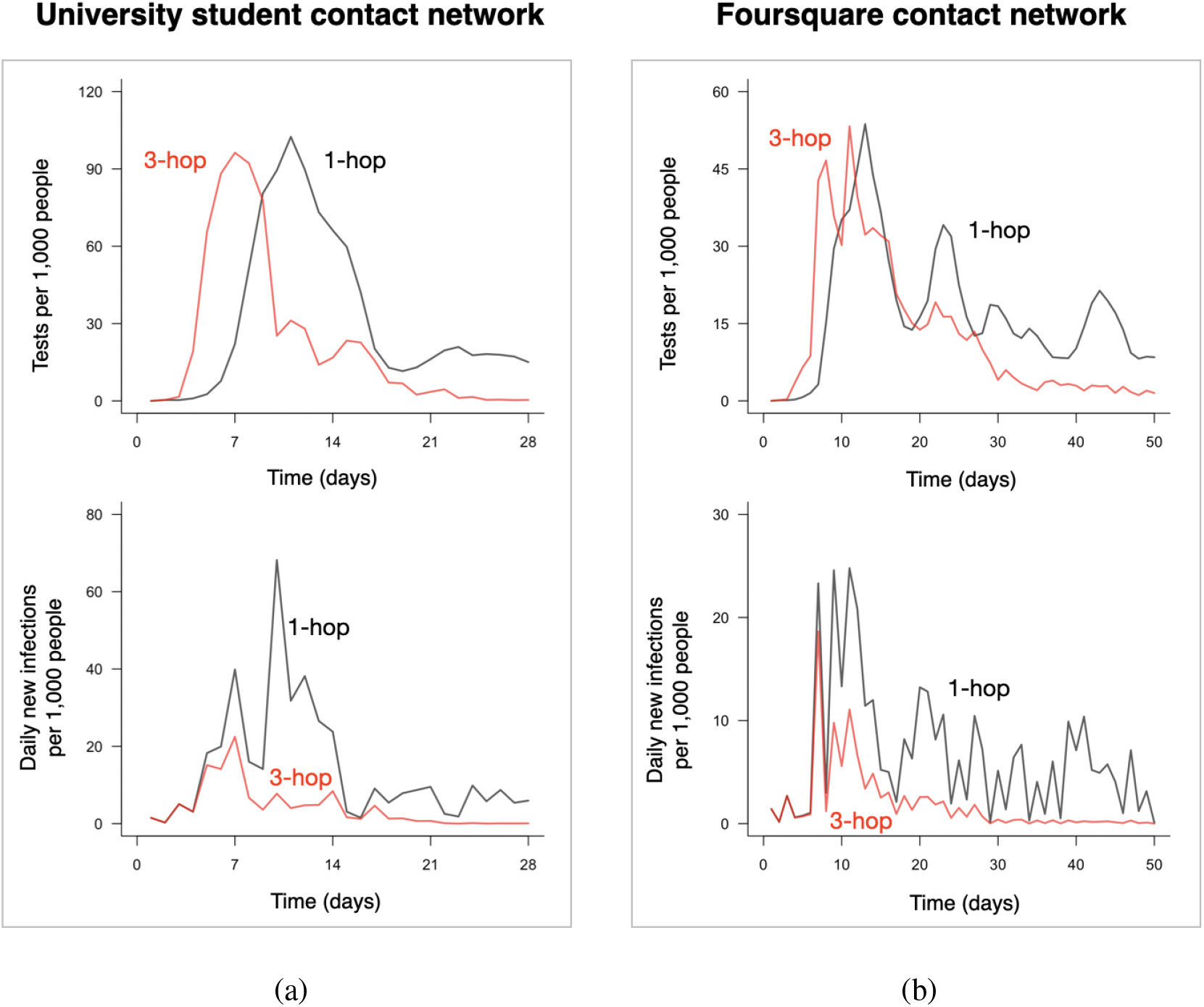
The number of tests over time and daily new infections over time for *University student contact network and Foursquare contact network*. The plots compare the numbers for 1-hop and *k*-hop contact tracings when *k* equals the corresponding threshold value. The first and the second column have been obtained for the same setups as the first column in Figure 3a and 4a, respectively.

**Figure 6:**
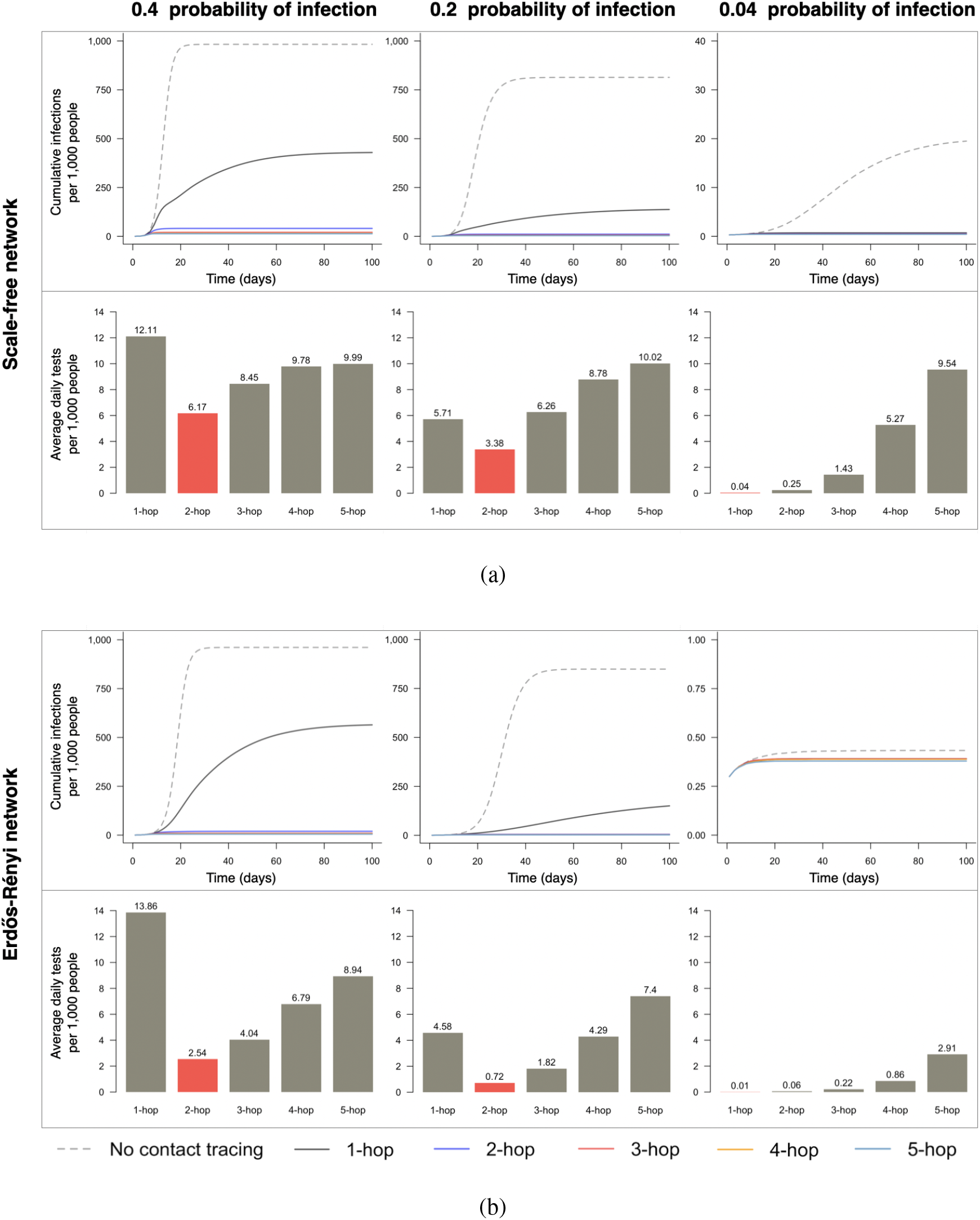
The cumulative number of infections and average daily tests required for *k*-hop contact tracing for various values of *k* for *synthetic networks*. The red colored bar corresponds to the threshold value for *k*. For *k* exceeding the threshold value, the curves for the cumulative number of infections heavily overlap and become indistinguishable.

We start with contact networks obtained from real-world data. We first consider the highest probability of infection (i.e., 0.4) and lowest cooperativity (i.e., 0.2) from our range of choices for the University student contact network. Recall that probability of infection and cooperativity are determined by human behavior and their respective increase and decrease enhance the spread of the disease and lower the reach of testing. Thus, this scenario represents a combination of elements of human behavior that render a populace most vulnerable to a pandemic (among our parameter choices). As *k* increases from 0 to 3, the cumulative number of infections considerably decreases, and subsequently decreases only marginally with further increase in *k*. 3-hop contact tracing achieves 80% reduction in the cumulative number of infections compared to 0-hop contacts tracing (i.e., no contact tracing), while 1-hop contact tracing achieves only 30% reduction (top left of Figure 3a). The total number of tests for *k*-hop contact tracing steadily decreases with increase in *k* from *k* = 1 to *k* = 3, and subsequently steadily increases as *k* increases further. This number is minimized for 3-hop contact tracing, at which value 27% fewer tests are required, overall, than 1-hop tracing (bottom left of Figure 3a). Thus, the threshold value is *k* = 3. Studying how the daily number of tests varies with time, it is somewhat higher initially for 3-hop contact tracing, but rapidly declines soon, as compared to 1-hop contact tracing (top of Figure 5a), leading to overall fewer number of tests for multi-hop contact tracing. These observations may be explained as follows. Multi-hop contact tracing may test greater number of individuals in early stages because it traces up to more hops even from the same number of confirmed cases (top of Figure 5a). However, this aggressive preemptive testing from early stages can rapidly mitigate the spread of infection earlier than 1-hop contact tracing through early identification and isolation of the infected (bottom of Figure 5a), thus fewer number of individuals transmit the virus and need tests with passage of time. The latter phenomenon more than compensates for the larger number of tests required in early stage for multi-hop contacts of those who test positive.

The above phenomena also recurs in the other contact network we obtain from real-world data, namely the Foursquare contact network. Figures 4 and 5b for the Foursquare contact network show identical trend as Figures 3 and 5a respectively for the University student contact network. For the same probability of infection (i.e., 0.4) and cooperativity (i.e., 0.2), the threshold is again *k* = 3. Again, the cumulative number of infections considerably decreases, as *k* increases from 0 to the threshold point of 3, and with subsequent increase in *k* the decrease in the cumulative number of infections is only marginal. 3-hop contact tracing achieves 81% reduction in the cumulative number of infections compared to 0-hop contact tracing, while 1-hop contact tracing achieves only 24% reduction (top left of Figure 4a). 37% fewer tests are required, overall, for 3-hop contact tracing as compared to 1-hop tracing (bottom left of Figure 4a). Again, although as compared to 1-hop contact tracing, 3-hop contact tracing initially needs somewhat higher number of daily tests, the number of daily tests in 3-hop contact tracing rapidly declines soon (top of Figure 5b), leading to overall fewer number of tests for 3-hop contact tracing. The higher number of tests in the early stages helps contain the spread of infection rapidly (bottom of Figure 5b).

All the phenomena reported above is replicated for other parameters for both the University student and Foursquare contact networks (other subfigures of Figure 3 and Figure 4), but the extent of the advantage and values of the thresholds differ. The differences in the specific numbers reveal that multi-hop contact tracing is more *resilient* to components of human behavior that enhance the spread of the disease and lower the reach of testing:

- When cooperativity increases to 1, the reach of testing is higher, and the efficacy of 1-hop tracing increases substantially. In University student contact network, it reduces cumulative infections by 80% compared to 0-hop tracing (top right of Figure 3a). In Foursquare contact network, it reduces cumulative infections by 83% compared to 0-hop tracing (top right of Figure 4a). Comparing the results for cooperativities of 1 and lower, we note that multi-hop contact tracing can offset the limitation arising from the lack of available information on contacts. This is because decrease in cooperativity reduces the reach of tests while multi-hop contact tracing increases the reach.
- As probability of infection decreases, the disease spreads slower, and 1-hop contact tracing becomes more and more effective, and the threshold value generally decreases (or remains the same). For University student contact network, refer to Figure 3b for the intermediate value of 0.2 for the probability of infection, and to Figure 8 in Supplementary Information for the lowest value of 0.04. For Foursquare contact network, refer to Figure 4b for the intermediate value of 0.2 for the probability of infection, and to Figure 9 in Supplementary Information for the lowest value of 0.04. In other words, therefore, employing multi-hop contact tracing is more beneficial when probability of infection is high.

**Figure 7:**
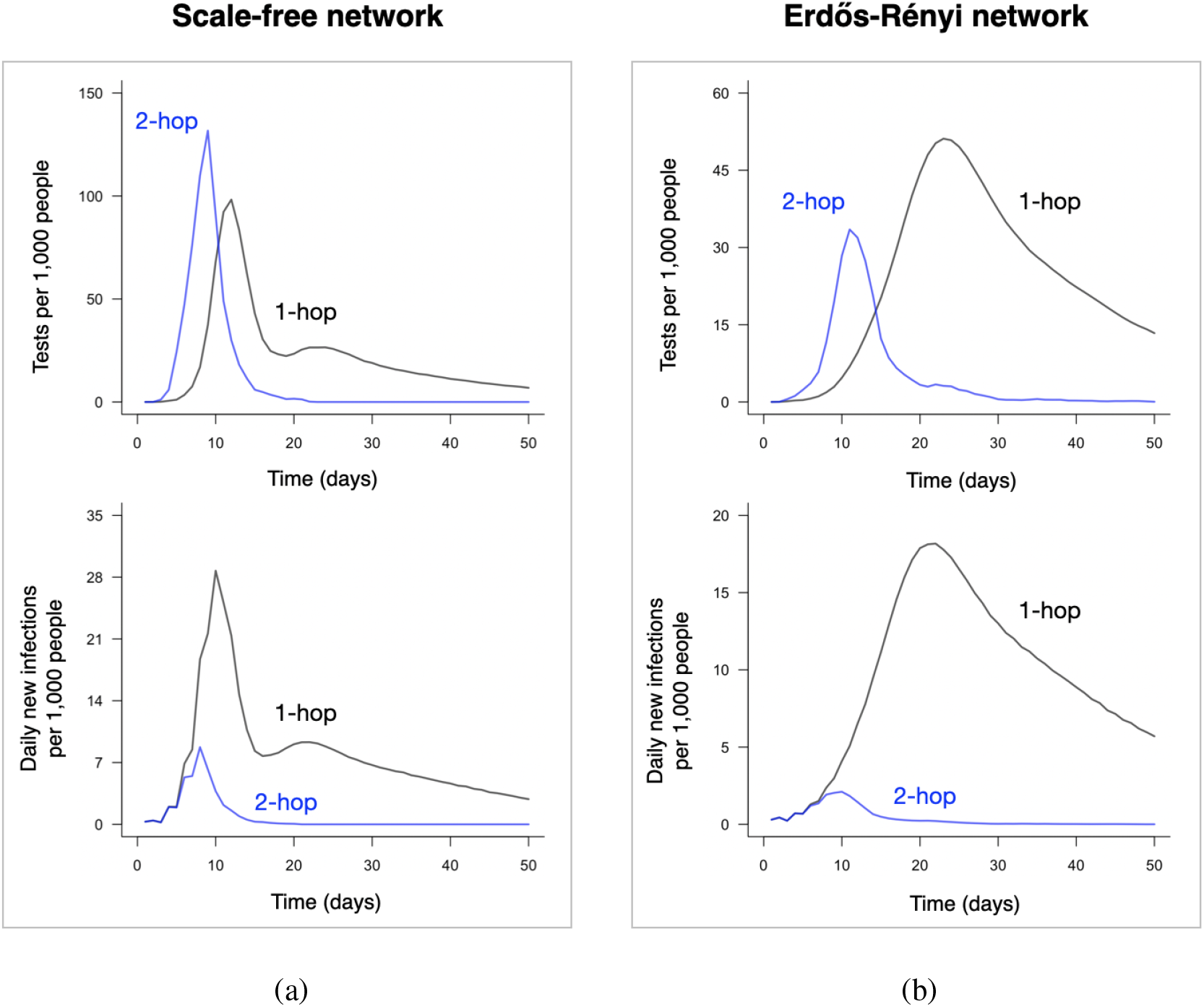
The number of tests over time and daily new infections over time for *synthetic networks*. The plots compare the numbers for 1-hop and *k*-hop contact tracings when *k* equals the corresponding threshold value. The first and the second column have been obtained for the same setups as the first column in Figure 6a and 6b, respectively.

We consider synthetic networks now (Figure 6). All the phenomena reported above is replicated, but the specifics differ. When probability of infection is 0.4, the threshold value is 2 for both the scale-free and Erdős Rényi networks (first column in Figure 6). In the two cases, despite the fact that the overall number of tests required in 2-hop contact tracing is significantly lower than that in 1-hop contact tracing, 2-hop contact tracing reduces the cumulative number of infections by 96% and 98%, respectively compared to 0-hop tracing, while 1-hop tracing reduces by 56% and 41%, respectively (top left of Figure 6a and 6b). The phenomenon of diminishing return is even more accentuated in these as the number of tests sharply increase with increase in *k* beyond *k* = 2 for *k*−hop contact tracing (bottom left of Figure 6a and 6b). Also, considering the variation of the number of tests over time, we notice that in scale-free networks, the number is significantly higher for multi-hop contact tracing than that for 1-hop contact tracing *initially* (top of Figure 7a). But, the decline in the number of tests required for multi-hop contact tracing becomes equally precipitous over time, and the rapid decline starts in a short time from the start of the testing period as well. Figure 6 plots the average of 1000 simulation runs over 1 realization of the synthetic topologies, Figure 11 in Supplementary Information reports the average of 100 runs over 10 realizations of the same synthetic topologies, corresponding plots in Figure 6 and Figure 11 show identical trend.

We now focus on the peak infection period and the average daily new infection count during this period for all the contact networks considered above. The duration of this period is a measure of how soon the infection is contained. The average daily new infection count during this period is a measure of the treatment-load health care centers experience in a critical period in which these have the potential to be overwhelmed. Under multi-hop contact tracing the peak infection period is shorter, in some cases considerably shorter, and the average daily new infection count during this period is invariably substantially lower, and therefore the healthcare facilities run lower risk of being overwhelmed. For example, in University student contact network and Foursquare contact network, considering the parameters as the first column in Figure 3a and 4a, respectively, for 3-hop contact tracing, the peak infection periods are 7 and 7 days, respectively, and an average of 8.8 (per 1000 population) and 3.6 (per 1000 population) daily new infections occurs during this period, respectively. On the other hand, for 1-hop contact tracing in these networks, the peak infection periods are 10 and 11 days, and an average of 18.6 (per 1000) and 8.6 (per 1000) daily new infections occurs during this period, respectively (Figure 5). Next, in scale-free and Erdős Rényi networks, considering the parameters as the first column in Figure 6, under 2-hop contact tracing, the peak infection periods are 8 and 10 days, respectively, and an average of 3 (per 1000) and 1.1 (per 1000) daily new infection occurs during this period, respectively. On the other hand, for 1-hop contact tracing in these topologies, the peak infection periods are 10 and 22 days, respectively, and an average of 8.9 (per 1000) and 7.7 (per 1000) daily new infections occurs during this period, respectively (Figure 7). Note that multi-hop contact tracing substantially reduces the peak infection period in Erdős Rényi network.

We now examine how increase in the latency period affects the results. In this case, the infected individuals become contagious later, thus, the potential of 1-hop contact tracing to detect and remove individuals before they become contagious or in the early period of their becoming contagious increases. Comparing the results for randomized latency periods of two different means, we note that this is indeed the case. In University student contact network, when the mean latency period is 1 day (2 days, respectively), 1-hop contact tracing can achieve 30% (43%, respectively) reduction in the cumulative number of infections compared to 0-hop contact tracing, and 3-hop contact tracing can achieve 80% (86%, respectively) reduction. In Foursquare contact network, when the mean latency period is 1 day (2 days, respectively), 1-hop contact tracing can achieve 24% (33%, respectively) reduction in the cumulative number of infections compared to 0-hop contact tracing, and 3-hop contact tracing can achieve 81% (86%, respectively) reduction. In scale-free network, when the mean latency period is 1 day (2 days, respectively), 1-hop contact tracing can achieve 56% (71%, respectively) reduction in the cumulative number of infections compared to 0-hop contact tracing, and 2-hop contact tracing can achieve 96% (98%, respectively) reduction. In Erdős Rényi network, when the mean latency period is 1 day (2 days, respectively), 1-hop contact tracing can achieve 41% (58%, respectively) reduction in the cumulative number of infections compared to 0-hop contact tracing, and 2-hop contact tracing can achieve 98% (99%, respectively) reduction. Refer to Figure 12 in Supplementary Information for a bar-graph representation of this data.

## Discussion

Our findings obtained through extensive simulations over a diverse set of four contact networks, starting from those obtained from real-world data to synthetic networks show that multi-hop contact tracing has the potential to substantially reduce total number of infections (which would in turn reduce fatalities and treatment load on healthcare facilities) and reduce overall testing load. It is more resilient to elements of human behavior that enhance the spread of the disease and lower the reach of testing. It also helps contain COVID-19 outbreaks within shorter times and reduces average daily new infection counts, specifically in the period up to when the daily new infection count peaks, which would in turn reduce the treatment load on healthcare facilities during the peak period. All these collectively have the potential to contain the outbreak without extensive lockdowns and economic downturns.

Our findings lead to the following recommendations for public health authorities:

- Deploy multi-hop contact tracing; usually testing up to secondary or tertiary contacts of those who test positive suffices.
- The recommended number of hops are on the higher end of the above range for lower latency periods and when human behavior enhances the spread of the disease and lowers the reach of testing, that is: 1) the probability of infection in each contact is high (e.g., from indoor contacts, longer durations of contacts, lack of protective gears and personal hygiene) 2) cooperativity is low. Given the imponderables in human behavior, the probability of infection and cooperativity can not usually be accurately estimated apriori. Thus, it is safer to err on the side of caution and opt for the higher end of the recommended range on the number of hops.
- Create the infrastructure for handling a larger testing load for limited periods, which will lead to a reduction in the overall testing load over time with the daily testing load expected to decline shortly after the testing starts (if multi-hop contact tracing is deployed).

Finally, multi-hop contact tracing may provide similar benefits for other infectious diseases which exhibit silent propagation (infection from individuals who do not show symptoms).

Note that the contact tracing recommendation of the CDC involves both testing and quarantining (even when the test result is negative) the direct or primary contacts of those who test positive. Along the same lines, recent works considered tracing and quarantining (or testing) the direct contacts of those who tested positive [1, 6, 9, 13]. Another recent work considered tracing and quarantining both primary and secondary contacts of those who test positive, and found that quarantining secondary contacts decreases the cumulative infection count compared to quarantining only the primary contacts, but also requires substantially higher number of quarantines [10]; [10] appears to reject the notion of quarantining secondary contacts and did not therefore explore quarantining tertiary or even more distant contacts. It does not investigate testing the primary and secondary contacts, perhaps because they would be quarantined regardless of the test results. We instead explore tracing and testing multi-hop contacts, testing eliminates the need for extensive quarantining as only those who test positive need to be isolated. Our results show that multi-hop contact tracing even reduces the number of tests required. The difference between our finding and the recent work arises because quarantine is a cumulative process in which each contact is quarantined for several days, while tests related to each positive patient are done only once. As to the primary contacts of those who test positive, we take no position on whether they should be quarantined even when they test negative, and leave that to the policies of the relevant public health authorities.

We now discuss the scenarios in which some of our assumptions may not hold. Tracing contacts and scheduling tests for those traced may require more than a day when individuals do not use contact tracing apps. PCR test results are not obtained same day if the testing site and laboratory are not co-located, though this delay may not affect the broad nature of our findings if those tested quarantine until the test results are known. Antigen tests give results in an hour but some of them reportedly record non-negligible proportion of false negatives. Depending on classifiers such as duration, environment (indoor or outdoor), usage of protective gears, observance of personal hygiene, different contacts may pass on infection with different probabilities. Assuming that such a probability is identical for all contacts with same infectious categories, which is what we did, is equivalent to considering an average over all contacts. Explicitly investigating the impact of 1) delay in tracing contacts and obtaining test results 2) errors in test results 3) non-uniform infection probabilities constitute directions for future research.

## Methods

### Construction of contact networks from real-world data

Our goal has been to evaluate the multi-hop contact tracing strategy using publicly-available data of human contact patterns. For evaluating the impact of multi-hop contact tracing, data sets need to involve large population sizes, otherwise length of the contact chains will be limited by the size of the target populace. Also, in reality, pandemic spread involves large target populaces and over several days, weeks and months. Data of human contact patterns is not plentiful in the public domain due to privacy and other concerns, the availability becomes even less for contact patterns of even moderate population sizes and over moderate periods of times (e.g., even several weeks). Nonetheless, we construct two different contact networks from data of real-world spatio-temporal records of human presence over certain periods of time: (1) University student contact network and (2) Foursquare contact network.

For the University student contact network, we utilize data collected by smartphones of University students, as part of the Copenhagen Networks Study [22]. The smartphones were equipped with Bluetooth cards which recorded proximity between participating students at 5-minute resolution. According to the definition of *close contact* by CDC [30], we only used proximity events between individuals that lasted more than 15 minutes in a row in the same day to construct daily contact networks. We postulated that two individuals had a contact if there are at least three consecutive proximity events with a 5-minute resolution between them. The constructed contact network has 2959.82 daily contacts on average among 672 individuals spanning 28 days. The strength of this data set is that it provides actual proximity events of a moderate number of individuals over several weeks (a moderate period of time).

For the Foursquare contact network, we utilized the data that users of Foursquare service (a Location-Based Social Networking or LBSN) made available in Social Media about their locations in Tokyo, Japan along with time-stamps. This dataset contains advertised locations (or checkins) collected for about 10 months (from 12 April 2012 to 16 February 2013). Each check-in contains information about the time at which the user visited the location, the GPS coordinates of the locations, and the nature of the locations (e.g., coffee shops, restaurants etc.) [32]. We use the first 100 days of data with at least one check-in. The strength of this data set is that it provides actual time-stamped locations of a large number of individuals, more than 2000, over several months (a long period of time). The weakness is that the contacts are still sparser than what arises in reality as the locations in question are usually crowded and much larger number of individuals actually visit these locations in overlapping time intervals but their whereabouts are not being reported in this dataset as they do not use this LBSN. The contact network will become denser if their presence can be considered. To compensate for this artificial sparsity we construct contact patterns based on the available data by postulating that people have had a contact if they have been at the same venue in the same day. Note that in reality individuals who have been at the same location in the same day do not always do so at the same time and are therefore not always in physical proximity to pass on the disease from one to another. Thus, the contact pattern we consider is denser than what the dataset actually provides, which may compensate for the artificial sparsity in question. The constructed contact network of Foursquare users has 5553.71 daily interaction on average among 2120 individuals.

The University student network involves spatio-temporal records of a smaller number of individuals and over a shorter period of time as compared to the Foursquare contact network. The contacts in the former are however based on actual proximity, while those in the latter are based on visits to the same location during the same day. Thus these two networks have complementary strengths and weaknesses. The evaluations can be repeated on more accurate and expansive contact patterns, which would combine the strengths of the two contact networks above, as they become available through collective efforts and enrichment of existing data repositories.

Finally during the contacts in these two contact networks a contagious individual passes the disease on to a susceptible individual with a certain probability (we mention how we choose these probabilities where we provide details on the Compartmental model of virus transmission and in Table 1 in Supplementary Information).

### Synthetic networks

The synthetic topologies consist of scale-free network [2, 3] and Erdős Rényi random network [3, 8]. Figure 6 have been provided for only one realization of scale-free network and Erdős Rényi random network, Figure 11 in Supplementary Information however shows plots of averages over 10 realizations of each.

The scale-free network topologies are generated by Barabási-Albert method where new nodes are added at each time step with *m* links that connect to existing nodes with a probability being proportional to the degree of the existing nodes; we set *m* = 2 to generate the network. We consider topologies of 10, 000 nodes and 19, 997 edges, thus average degree of a node is ⟨*k*⟩ = 3.9994. For the single realization we use in Figure 6a, diameter and average path length are 9 and 5.01, respectively. Figure 10a in Supplementary Information shows that for the one realization we consider in Figure 6a, the degree distribution is well approximated by power-law with degree exponent 3, as should be for scale-free networks with the same degree exponent. Thus, the single realization in question is a typical scale-free network.

The Erdős Rényi network consists of 10, 000 nodes which are connected with 20, 000 randomly placed edges, thus the average degree of a node is ⟨*k*⟩ = 4. For the one realization we consider in Figure 6b, diameter (i.e., the greatest distance between pair of nodes of connected components) is 14 and average path length (i.e., average distance along the existing paths) is 6.76. Figure 10b in Supplementary Information shows that for the one realization we consider in Figure 6b, the degree distribution is well approximated by Poisson distribution with parameter ⟨*k*⟩, as should be for Erdős Rényi random networks. Thus, the single realization in question is a typical Erdős Rényi random network.

### Compartmental model of virus transmission

We use a *discrete time compartmental disease model* to model the progression of COVID-19 where the transition from each compartment to the next happens after a random amount of time with a geometric distribution. Compartmental model is also used in [1, 31]. Different stages of the disease are shown in Figure 2. The Compartmental model consists of the following stages: *Susceptible (S), Presymptomatic-Latent (I*_*p*_*-L), Presymptomatic (I*_*p*_*), Symptomatic (I*_*s*_*), Ready-to-Test (RT), Asymptomatic-Latent (I*_*a*_*-L), Asymptomatic (I*_*a*_*), Recovered (R)*, and *Dead (D)*. Only symptomatic individuals show symptoms, while presymptomatic, symptomatic and asymptomatic individuals infect others.

When a susceptible (*S*) individual comes into contact with a symptomatic (*I*_*s*_) individual, he is infected with the probability of infection *β*_*s*_. Similarly, a presymptomatic (*I*_*p*_) and an asymptomatic (*I*_*a*_) individual infects a susceptible upon contact with probabilities, *β*_*p*_(= *γ*_*p*_*β*_*s*_), *β*_*a*_(= *γ*_*a*_*β*_*s*_), respectively, where *γ*_*p*_ and *γ*_*a*_ are respectively infectiousness of presymptomatic and asymptomatic individuals relative to symptomatic individuals. If a susceptible individual *i* interacts with *l* symptomatic, *m* presymptomatic, and *n* asymptomatic individuals at time *t* − 1, the probability that the susceptible individual is infected at time *t* is 1 − (1 − *β*_*s*_)^*l*^(1 − *β*_*p*_)^*m*^(1 − *β*_*a*_)^*n*^.

Once an individual is infected he becomes contagious after a geometrically distributed latency time, whose statistics depends on whether he will develop symptoms at some point or otherwise. Following the nomenclature in compartmental models already utilized for COVID-19, we assume that an infected individual becomes asymptomatic-latent (with probability *p*_*a*_) or presymptomatic-latent (with probability 1 − *p*_*a*_). The asymptomatic-latent (*I*_*a*_-*L*) individuals never develop symptoms, do not infect others for an mean latency duration of 1/λ, and subsequently become contagious, at which stage we call them asymptomatic or *I*_*a*_ for simplicity. An asymptomatic individual remains contagious for a geometrically distributed random duration with mean 1/*r*_*a*_, after which he recovers. We now consider the other compartment an individual enters after infection, the presymptomatic-latent compartment. A presymptomatic-latent individual becomes contagious after a mean latency period of 1/λ, at which point we call him presymptomatic or *I*_*p*_. He remains presymptomatic for a geometrically distributed duration with mean 1/*α*; after this duration he develops symptoms and is called symptomatic. A symptomatic individual continues to infect his contacts until he opts for testing (*RT*). The duration for which a symptomatic individual infects others is geometrically distributed with mean 1/*w*. Once this duration ends, the patient quarantines himself and does not infect others. He ultimately dies (*D*) with probability *p*_*d*_, or recovers (*R*) with probability 1 − *p*_*d*_, after a geometrically distributed duration whose mean is 1/*r*_*s*_. We do not consider that individuals can be reinfected.

In all the networks, except in the University student contact network, we consider that initially all but three individuals are susceptible, among the three there is one each of presymptomatic, symptomatic and asymptomatic. Since the University student contact network involves fewer individuals, we consider a fewer number of initially infected individuals. Specifically we consider that initially all but one symptomatic individual are susceptible.

The parameters we choose and further justification for the choices are listed in Table 1 in Supplementary Information.

### Multi-hop contact tracing process

Every day individuals who are *ready to test* by virtue of showing symptoms are tested and isolated if they test positive. Considering an individual who tests positive on day *t*, we describe the process of tracing his *k*-hop contacts on day *t* and testing them on day *t*+1. On day *t* after the individual in question tests positive, the public health authority traces his *k*-hop contacts, over the last 14 days, and informs them that they may have been exposed. If cooperativity is *q*, only *q* proportion of interactions per day can be identified. Tests are scheduled on day *t*+1. Those scheduled to be tested are isolated from everyone else on the day of the test. The test results are available in the same day, and those who test positive are isolated until they recover and those who test negative can resume their normal activities. Individuals who test positive will not be tested again, but those who test negative can be tested again after 3 days from the test date if they have direct or indirect interactions with any one who tests positive or if they show symptoms.

## Data Availability

The raw data used to construct University student contact network was previously published in P. Sapiezynski, A. Stopczynski, D. D. Lassen, and S. Lehmann. Interaction data from the Copenhagen Networks Study. Scientific Data, 6(1):1-10, 2019. The raw data used to construct Foursquare contact network was previously published in Dingqi Yang, Daqing Zhang, Vincent W. Zheng, Zhiyong Yu. Modeling User Activity Preference by Leveraging User Spatial Temporal Characteristics in LBSNs. IEEE Trans. on Systems, Man, and Cybernetics, which is available at https://sites.google.com/site/yangdingqi/home/foursquare-dataset. All other data generated or analyzed during this study are available from the corresponding author upon request.

## Data availability

The raw data used to construct University student contact network was previously published in [22]. The raw data used to construct Foursquare contact network was previously published in [32], which is available at https://sites.google.com/site/yangdingqi/home/foursquare-dataset. All other data generated or analyzed during this study are available from the corresponding author upon request.

## Code availability

Custom code used to produce the results in this study is available from the corresponding author upon request.

## Supplementary Information

**Figure 8:**
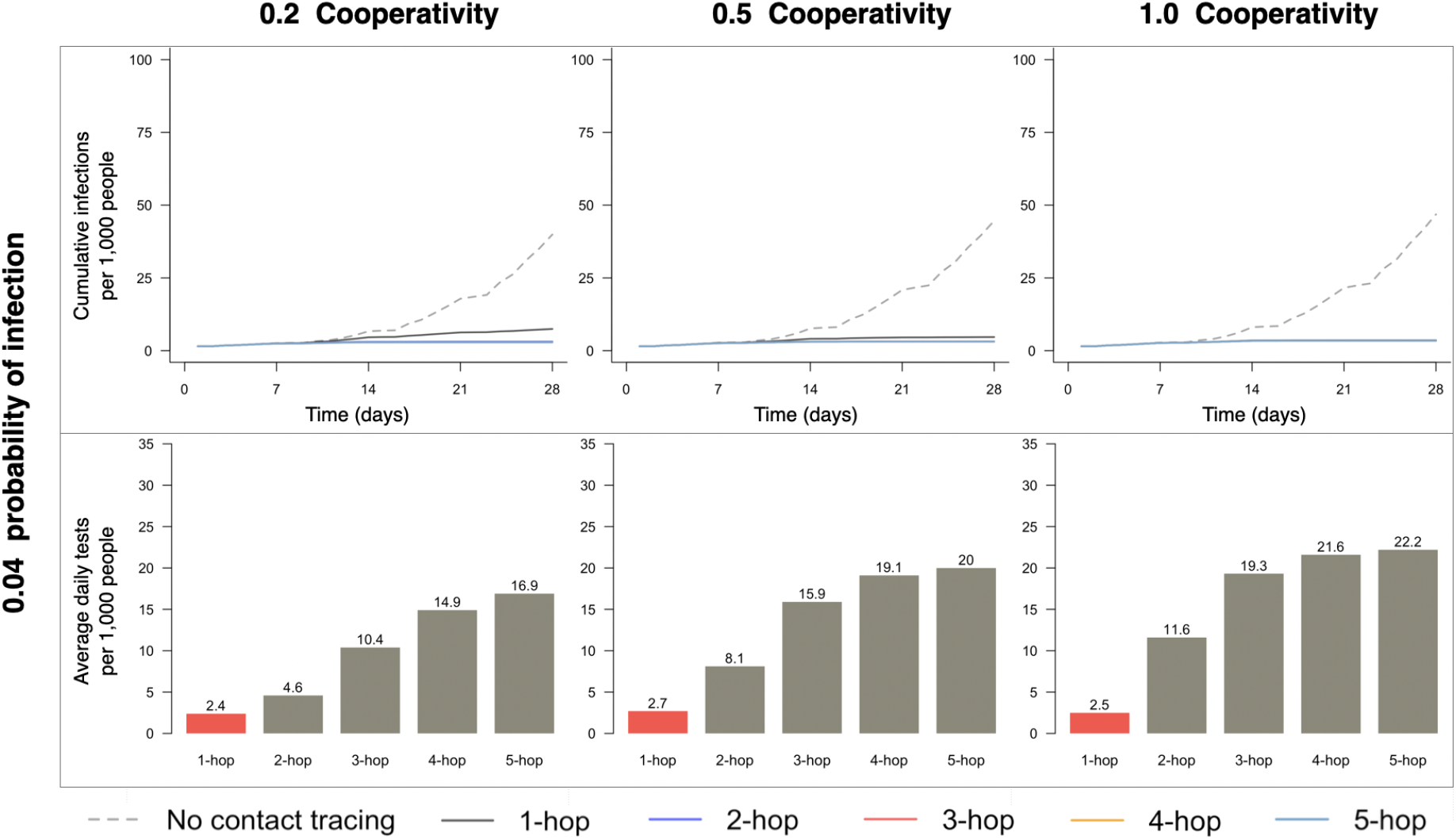
The cumulative number of infections and average daily tests required for *k*-hop contact tracing for various values of *k* for *University student contact network*. The probability of infection is 0.04. The red colored bar corresponds to the threshold value for *k*. For *k* exceeding the threshold value, the curves for the cumulative number of infections heavily overlap and become indistinguishable.

**Figure 9:**
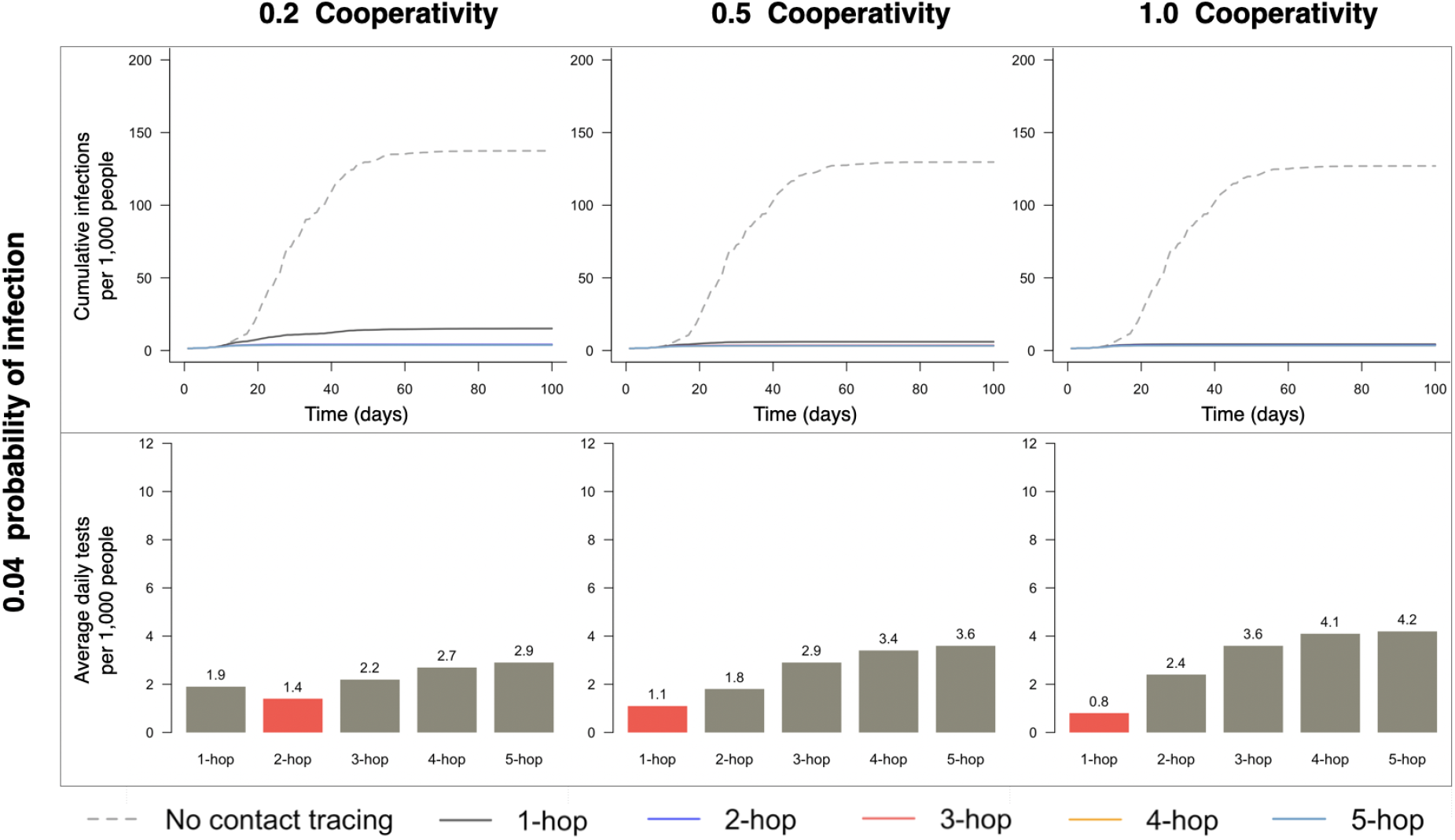
The cumulative number of infections and average daily tests required for *k*-hop contact tracing for various values of *k* for *Foursquare contact network*. The probability of infection is 0.04. The red colored bar corresponds to the threshold value for *k*. For *k* exceeding the threshold value, the curves for the cumulative number of infections heavily overlap and become indistinguishable.

**Figure 10:**
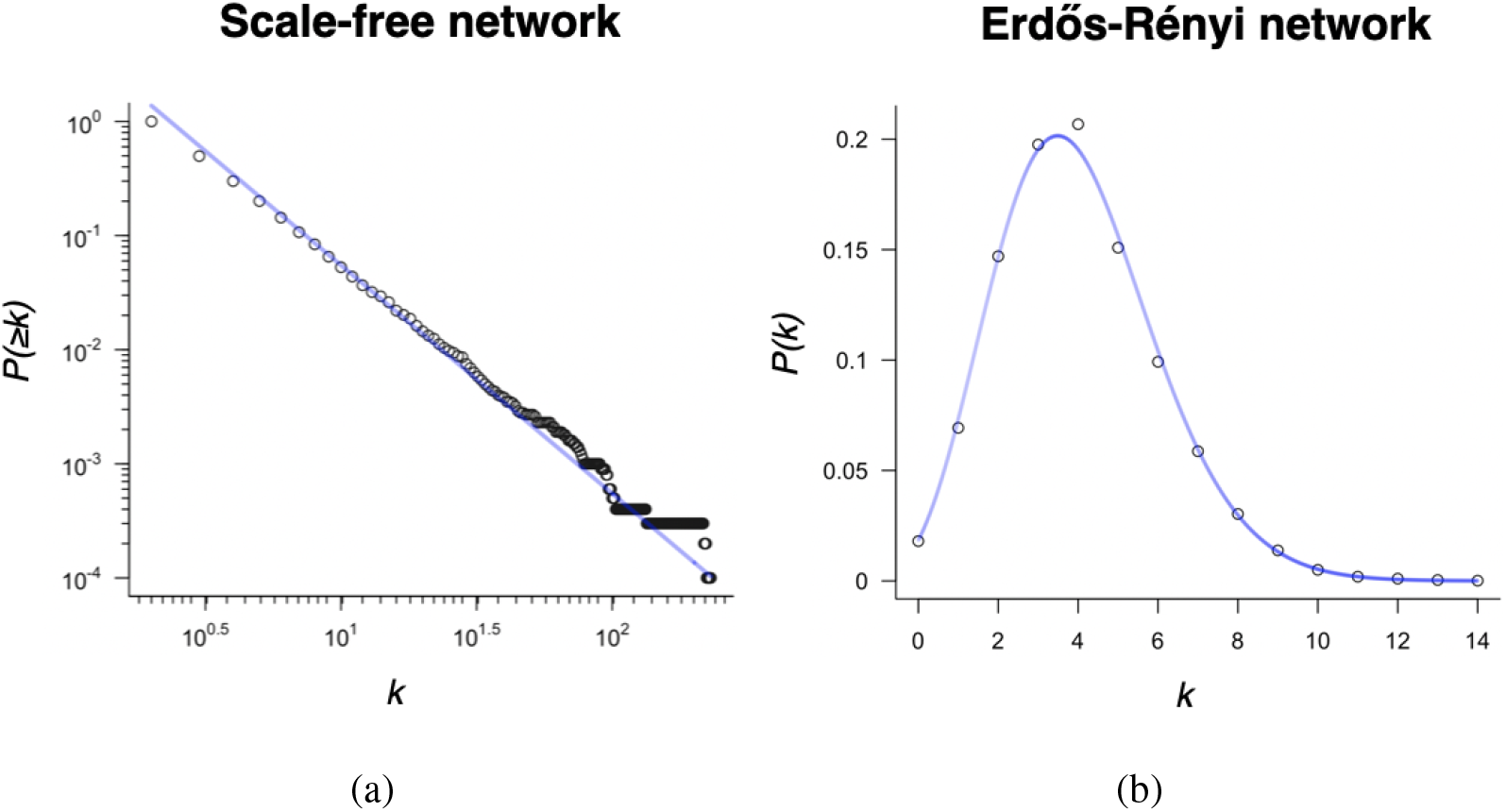
Degree distribution of the respective network realizations of the scale-free and Erdős Rényi network used for the data plotted in Figures 6 and 7. (a) The points represent the cumulative degree distribution for the scale-free network, and the slope of the line is 2 which is the theoretical exponent of this power-law distribution. Those are plotted on a double logarithmic scale. (b) The points represent the degree distribution for the Erdős Rényi network, and the line represents the Poisson distribution with parameter 4.

**Figure 11:**
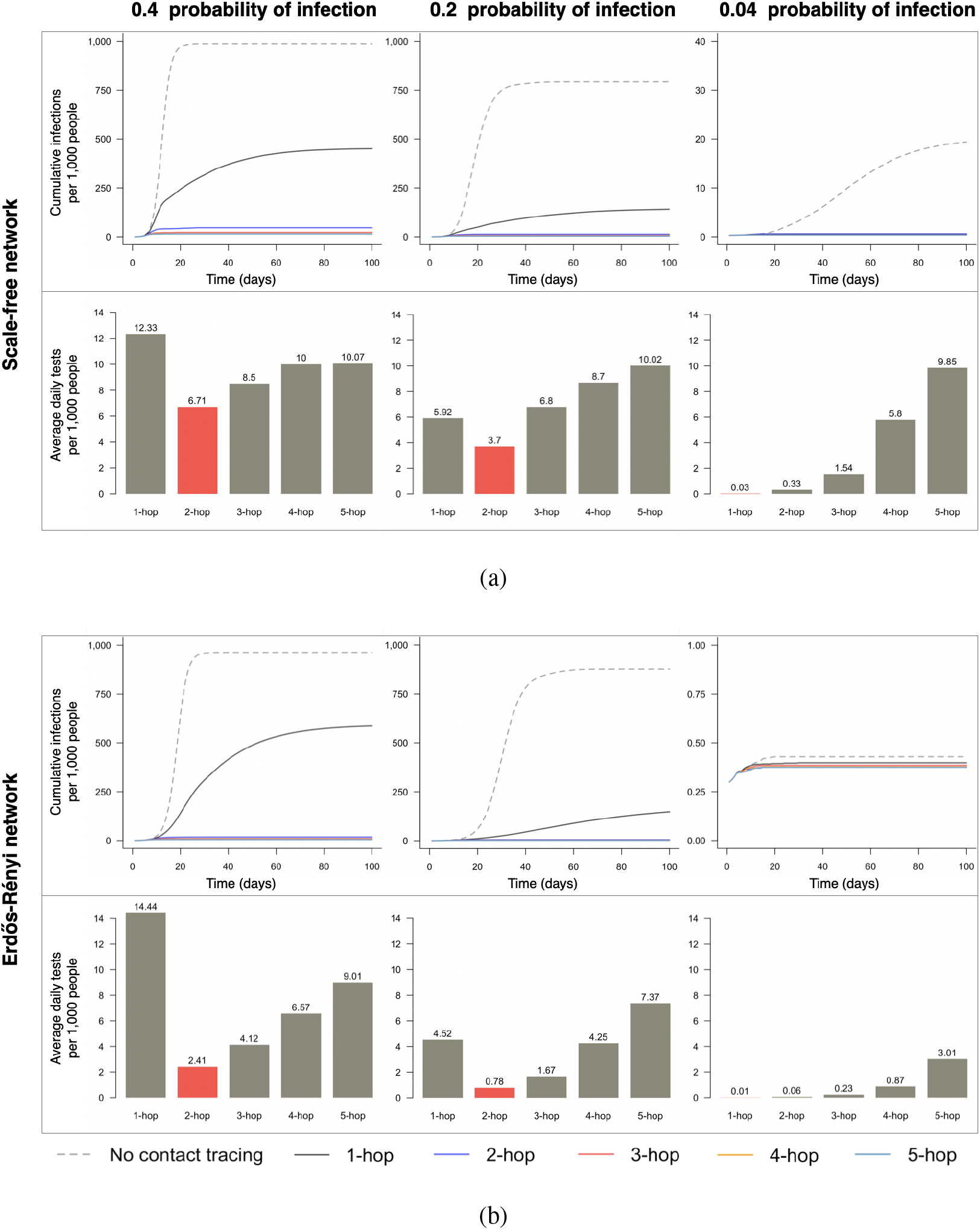
Cumulative infection count and total number of tests needed for multi-hop contact tracing policy. The data has been obtained for the same setups as in Figure 6, only difference is that this figure plots the results averaged over 10 network realizations and 100 simulation runs on each network realization, while Figure 6 plots the results averaged over 1000 simulation runs over 1 network realization. This figure closely resembles Figure 6 both in overall trends and specific values of the data points.

**Figure 12:**
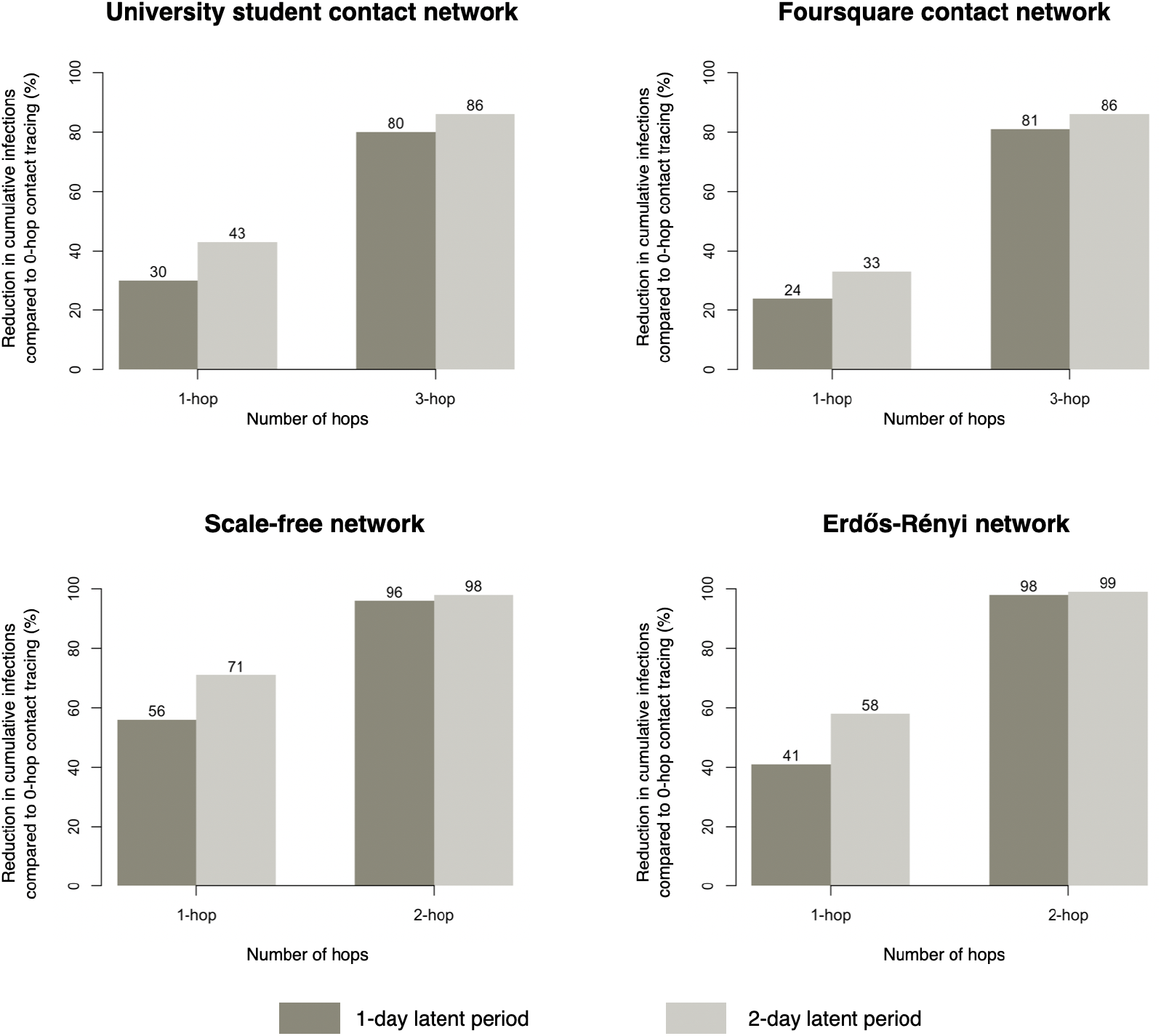
Comparison of cumulative infection for different values of mean latency period. We use probability of infection as 0.4. The cooperativities are 0.2 and 1 respectively for ***University student & Foursquare networks*** and static synthetic networks. The plots compare the numbers for 1-hop and *k*-hop contact tracings when *k* equals the corresponding threshold value.

**Table 1:** Values of disease parameters

| Parameter | Notation | Value | Reference & Description |
| --- | --- | --- | --- |
| Probability of infection<br>(Probability with which a symptomatic individual infects a susceptible in an interaction) | $\beta_s$ | 0.4, 0.2, 0.04 | Assumed various scenarios |
| Infectiousness of presymptomatic individuals relative to symptomatic | $\gamma_p$ | 1 | Inferred from [12] suggesting significant presymptomatic transmission |
| Infectiousness of asymptomatic individuals relative to symptomatic | $\gamma_a$ | 0.75 | Best estimate by [25] from prior studies [16],[34],[33],[18],[20] |
| Proportion of infections that are asymptomatic | $p_a$ | 0.4 | [25],[21] |
| Mean latency period <sup>†</sup> | $1/\lambda$ | 1, 2 days | Inferred from [19] |
| Mean duration in asymptomatic stage | $1/r_a$ | 7 days | Inferred from [5],[19] |
| Mean incubation period<br>(period between infection and the onset of symptoms) | $1/\lambda + 1/\alpha$ | 5 days | [17],[15] |
| Mean duration from symptom onset to testing | $1/w$ | 4 days | Inferred from [4] |
| Mean duration of symptom onset to recovery or death | $1/w + 1/r_s$ | 14 days | Inferred from [24], [5] |
| Fraction of symptomatics who die | $p_d$ | 0.0065 | [25] |

<sup>†</sup> All Figures use a mean latency period of 1, except Figure 12 in which we compare the results for mean latency periods of 1 and 2.

## References

1. A. Aleta, D. Martín-Corral, A. P. y Piontti, M. Ajelli, M. Litvinova, M. Chinazzi, N. E. Dean, M. E. Halloran, I. M. Longini Jr, S. Merler, et al. Modelling the impact of testing, contact tracing and household quarantine on second waves of COVID-19. Nature Human Behaviour, 4(9):964–971, 2020.

2. A.-L. Barabási and R. Albert. Emergence of scaling in random networks. Science, 286(5439): 509–512, 1999.

3. A.-L. Barabási et al. Network science. Cambridge university press, 2016.

4. M. Biggerstaff, M. A. Jhung, C. Reed, A. M. Fry, L. Balluz, and L. Finelli. Influenza-like illness, the time to seek healthcare, and influenza antiviral receipt during the 2010–2011 influenza season—United States. The Journal of infectious diseases, 210(4):535–544, 2014.

5. A. W. Byrne, D. McEvoy, A. B. Collins, K. Hunt, M. Casey, A. Barber, F. Butler, J. Griffin, E. A. Lane, C. McAloon, K. O’Brien, P. Wall, K. A. Walsh, and S. J. More. Inferred duration of infectious period of SARS-CoV-2: rapid scoping review and analysis of available evidence for asymptomatic and symptomatic COVID-19 cases. BMJ Open, 10(8), 2020. ISSN 2044-6055. doi: 10.1136/bmjopen-2020-039856. URL https://bmjopen.bmj.com/content/10/8/e039856.

6. W. A. Chiu, R. Fischer, and M. L. Ndeffo-Mbah. State-level needs for social distancing and contact tracing to contain COVID-19 in the United States. Nature Human Behaviour, 2020.

7. D. K. Chu, E. A. Akl, S. Duda, K. Solo, S. Yaacoub, H. J. Schünemann, A. El-harakeh, A. Bognanni, T. Lotfi, M. Loeb, et al. Physical distancing, face masks, and eye protection to prevent person-to-person transmission of sars-cov-2 and covid-19: a systematic review and meta-analysis. The Lancet, 2020.

8. P. Erdős and A. Rényi. On the evolution of random graphs. Publ. Math. Inst. Hung. Acad. Sci, 5(1):17–60, 1960.

9. L. Ferretti, C. Wymant, M. Kendall, L. Zhao, A. Nurtay, L. Abeler-Dörner, M. Parker, D. Bonsall, and C. Fraser. Quantifying SARS-CoV-2 transmission suggests epidemic control with digital contact tracing. Science, 368(6491), 2020.

10. J. A. Firth, J. Hellewell, P. Klepac, S. Kissler, A. J. Kucharski, and L. G. Spurgin. Using a real-world network to model localized COVID-19 control strategies. Nature Medicine, pages 1–7, 2020.

11. G. Guglielmi. The explosion of new coronavirus tests that could help to end the pandemic. Nature, 2020. https://www.nature.com/articles/d41586-020-02140-8.

12. X. He, E. H. Lau, P. Wu, X. Deng, J. Wang, X. Hao, Y. C. Lau, J. Y. Wong, Y. Guan, X. Tan, et al. Temporal dynamics in viral shedding and transmissibility of COVID-19. Nature medicine, 26(5):672–675, 2020.

13. J. Hellewell, S. Abbott, A. Gimma, N. I. Bosse, C. I. Jarvis, T. W. Russell, J. D. Munday, A. J. Kucharski, W. J. Edmunds, F. Sun, et al. Feasibility of controlling COVID-19 outbreaks by isolation of cases and contacts. The Lancet Global Health, 2020.

14. R. R. Lash, C. V. Donovan, A. T. Fleischauer, Z. S. Moore, G. Harris, S. Hayes, M. Sullivan, A. Wilburn, J. Ong, D. Wright, et al. COVID-19 Contact Tracing in Two Counties-North Carolina, June-July 2020. MMWR. Morbidity and mortality weekly report, 69(38):1360–1363, 2020.

15. S. A. Lauer, K. H. Grantz, Q. Bi, F. K. Jones, Q. Zheng, H. R. Meredith, A. S. Azman, N. G. Reich, and J. Lessler. The incubation period of coronavirus disease 2019 (covid-19) from publicly reported confirmed cases: estimation and application. Annals of internal medicine, 172(9):577–582, 2020.

16. S. Lee, T. Kim, E. Lee, C. Lee, H. Kim, H. Rhee, S. Y. Park, H.-J. Son, S. Yu, J. W. Park, et al. Clinical course and molecular viral shedding among asymptomatic and symptomatic patients with SARS-CoV-2 infection in a community treatment center in the republic of korea. JAMA Internal Medicine, 2020.

17. N. M. Linton, T. Kobayashi, Y. Yang, K. Hayashi, A. R. Akhmetzhanov, S.-m. Jung, B. Yuan, R. Kinoshita, and H. Nishiura. Incubation period and other epidemiological characteristics of 2019 novel coronavirus infections with right truncation: a statistical analysis of publicly available case data. Journal of clinical medicine, 9(2):538, 2020.

18. Y. Liu, L.-M. Yan, L. Wan, T.-X. Xiang, A. Le, J.-M. Liu, M. Peiris, L. L. Poon, and W. Zhang. Viral dynamics in mild and severe cases of covid-19. The Lancet Infectious Diseases, 2020.

19. S. Ma, J. Zhang, M. Zeng, Q. Yun, W. Guo, Y. Zheng, S. Zhao, M. H. Wang, and Z. Yang. Epi-demiological parameters of coronavirus disease 2019: a pooled analysis of publicly reported individual data of 1155 cases from seven countries. Medrxiv, 2020.

20. J. Y. Noh, J. G. Yoon, H. Seong, W. S. Choi, J. W. Sohn, H. J. Cheong, W. J. Kim, and J. Y. Song. Asymptomatic infection and atypical manifestations of covid-19: comparison of viral shedding duration. The Journal of Infection, 2020.

21. D. P. Oran and E. J. Topol. Prevalence of asymptomatic SARS-CoV-2 infection: A narrative review. Annals of Internal Medicine, 2020.

22. P. Sapiezynski, A. Stopczynski, D. D. Lassen, and S. Lehmann. Interaction data from the Copenhagen Networks Study. Scientific Data, 6(1):1–10, 2019.

23. R. Service. In ‘milestone,’ fda oks simple, accurate coronavirus test that could cost just $5. Science, 2020. https://www.sciencemag.org/news/2020/08/milestone-fda-oks-simple-accurate-coronavirus-test-could-cost-just-5.

24. The U.S. Centers for Disease Control and Prevention (CDC). COVID-19 pandemic planning scenarios. May 2020. https://www.cdc.gov/coronavirus/2019-ncov/hcp/planning-scenarios-archive/planning-scenarios-2020-05-20.pdf.

25. The U.S. Centers for Disease Control and Prevention (CDC). COVID-19 pandemic planning scenarios. Aug. 2020. https://www.cdc.gov/coronavirus/2019-ncov/hcp/planning-scenarios.html#definitions.

26. The U.S. Centers for Disease Control and Prevention (CDC). Scientific brief: SARS-CoV-2 and potential airborne transmission. Oct. 2020. https://www.cdc.gov/coronavirus/2019-ncov/more/scientific-brief-sars-cov-2.html.

27. The U.S. Centers for Disease Control and Prevention (CDC). Public health guidance for community-related exposure. July 2020. https://www.cdc.gov/coronavirus/2019-ncov/php/public-health-recommendations.html.

28. The U.S. Centers for Disease Control and Prevention (CDC). How COVID-19 spreads. Oct. 2020. https://www.cdc.gov/coronavirus/2019-ncov/prevent-getting-sick/how-covid-spreads.html.

29. The U.S. Centers for Disease Control and Prevention (CDC). Overview of testing for SARS-CoV-2 (COVID-19). Sept. 2020. https://www.cdc.gov/coronavirus/2019-ncov/hcp/testing-overview.html.

30. The U.S. Centers for Disease Control and Prevention (CDC). Contact tracing for COVID-19. Sept. 2020. https://www.cdc.gov/coronavirus/2019-ncov/php/contact-tracing/contact-tracing-plan/contact-tracing.html.

31. C. J. Worby and H.-H. Chang. Face mask use in the general population and optimal resource allocation during the COVID-19 pandemic. medRxiv, 2020.

32. D. Yang, D. Zhang, V. W. Zheng, and Z. Yu. Modeling user activity preference by leveraging user spatial temporal characteristics in LBSNs. IEEE Transactions on Systems, Man, and Cybernetics: Systems, 45(1):129–142, 2014.

33. R. Zhou, F. Li, F. Chen, H. Liu, J. Zheng, C. Lei, and X. Wu. Viral dynamics in asymptomatic patients with covid-19. International Journal of Infectious Diseases, 2020.

34. L. Zou, F. Ruan, M. Huang, L. Liang, H. Huang, Z. Hong, J. Yu, M. Kang, Y. Song, J. Xia, et al. SARS-CoV-2 viral load in upper respiratory specimens of infected patients. New England Journal of Medicine, 382(12):1177–1179, 2020.

